# PVP1–The People’s Ventilator Project: A fully open, low-cost, pressure-controlled ventilator

**DOI:** 10.1101/2020.10.02.20206037

**Authors:** Julienne LaChance, Tom J. Zajdel, Manuel Schottdorf, Jonny L. Saunders, Sophie Dvali, Chase Marshall, Lorenzo Seirup, Daniel A. Notterman, Daniel J. Cohen

## Abstract

We present a fully open ventilator platform–The People’s Ventilator: PVP1– with complete documentation and detailed build instructions, and a DIY cost of $1,300 USD. Here, we validate PVP1 against key performance criteria specified in the U.S. Food and Drug Administration’s Emergency Use Authorization for Ventilators. Notably, PVP1 performs well over a wide range of test conditions and has been demonstrated to perform stably for a minimum of 72,000 breath cycles over three days with a mechanical test lung. As an open project, PVP1 can enable both future educational, academic, and clinical developments in the ventilator space.

## Introduction

### Access to software and hardware

Key resources for the People’s Ventilator Project (PVP1) are available online

- Source Code and Community
- Documentation
- PyPi - `pip install pvp`

### Ventilator needs in the 21st century

The global COVID-19 pandemic has highlighted the need for a low-cost, rapidly-deployable ventilator solution for the current and future pandemics. While safe and robust ventilation technology exists in the commercial sector, there exist a small number of suppliers who have been unable to meet the extreme demands for ventilators during a pandemic. Moreover, the specialized and proprietary equipment developed by medical device manufacturers can be prohibitively expensive and inaccessible in low-resource areas (1). Moreover, ventilation as a technology is needed globally beyond pandemics for applications spanning neonatal intensive care, surgical anaesthesia, life support, and general respiratory treatments. Hence there is a clear need for a broader range of solutions, both for research and clinical applications.

In response to these challenges, we present a fully open-source, rapid-deploy ventilator design with minimal reliance on specialized medical devices and manufacturing equipment. The People’s Ventilator Project (PVP1) is a pressure-controlled, fully automatic mechanical ventilator that can be built for less than $1,300 by a single person in few days, requiring neither specialized tools nor specialist knowledge. As a point of reference, the lower-end average market values of open ventilators (such as the freely-released Puritan Bennett 560 (Medtronic, Inc.) (2) or the Mechanical Ventilator Milano (Elemaster, Inc.) (3)) cost approximately $10,000. PVP1’s parts were selected for widespread availability, and its modular software was designed to support component substitutions and extensions to new ventilation modes, thereby increasing global access to critical-care ventilation technology.

Open source medical technology can improve the capability and access to medical technology as a whole in several ways: (1) enabling faster device innovation with lower costs (4); (2) increasing economic value, with associated public benefits, compared to traditional proprietary development (5); (3) facilitating external review and inspection by avoiding black-box hardware and software designs, and (4) providing a benchmark for innovation towards next-generation technology such as smart ventilators (6). Finally, the open source approach can make these problems more accessible to academic researchers, thereby greatly expanding the ability to train students in approaching such problems as well as encouraging unconventional approaches. While many pandemic ventilator projects began as open-source initiatives, these often transitioned to a closed format due to the strong structural and regulatory incentives to enter into industrial partnerships. Our goal with PVP1 was to provide a completely open build guide and software platform for a functional, pressure-controlled ventilator designed for FDA Emergency Use Authorization standards.

### Design Philosophy and System Capabilities

PVP1 is an automated ventilator that natively supports pressure-control ventilation (PCV), spontaneous inhalation monitoring ventilation (SIMV), and key alarms specified by regulatory agencies (e.g. high airway pressure, etc.). Pressure control was chosen over volume control because it is known to be safer (7) with respect to barotrauma risk, and SIMV was implemented because it increases the range of patients and conditions for which PVP1 can be used. Summarized in Fig. 1A, PVP1 operates as a computer-controlled, timed-cycle ventilator that requires only medical air and the patient-side respiratory tubing to be operated. To date, PVP1 has been set up three times by two different teams and run continuously for over 300 hours with no alarms or failures noted (representative data from a physical test-lung shown in Fig. 1D).

**Fig. 1.**
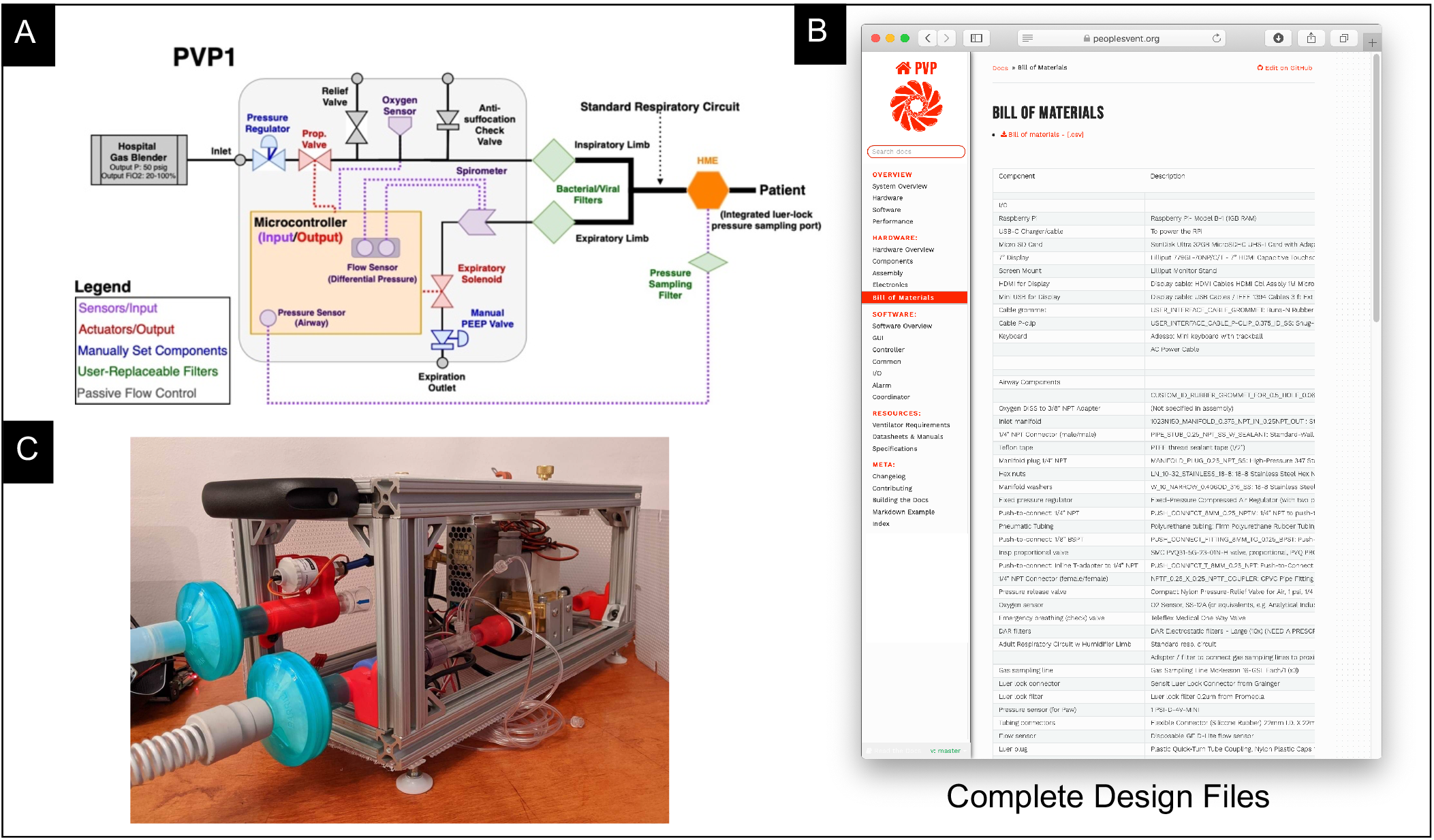
A system overview of the People’s Ventilator Project. A) overview of respiratory circuit. B) A snapshot of the online documentation demonstrating a complete web-portal optimized for sharing documentation and build instructions. PVP1 Bill of Materials is highlighted. C) The assembled device.

PVP1 was designed from the ground up to actually be a fully open source and transparent ventilation system, and as such we have made the entirety of our source code, electronics, bill-of-materials, complete CAD assembly, testing results, and relevant schematics open to the public (Fig. 1B). PVP1 follows the best practices for open projects (8): (i) transparent and public communication on our github repository, (ii) standardized and automated tests of individual modules and the full system with complete coverage using Travis CI (9), (iii) merger of pull requests only after passed tests and independent code review, (iv) high-level hardware and build documentation, and API-level documentation generated from docstrings and (v) PVP1 made accessible via pip and the Python Package Index which allows anyone to easily install, run, and experiment with the software.

We feel strongly that open design should include justification of design decisions and should be able to serve as a teaching and learning tool. As such, our Supporting Information here and our full documentation not only enable someone to build PVP1, but to learn why and how we chose each key component. Based on independent validation from collaborators, following our guidelines will lead to a functional ventilator in less than 3 days of hands-on-work. To further mitigate risk and expedite exploration and evaluation of the PVP1 platform, we provide the ability to run a complete simulation of the PVP1 system on any computer.

PVP1 was specifically designed for the constraints of a pandemic such as COVID-19, and special care was chosen to specify reliable, commercial, off-the-shelf components outside of the traditional ventilator or scientific supply chains. All components are available globally from general hardware suppliers anthe chosen parts did not experience supply chain disruptions due to COVID during the period of development. The internal layout and chassis design are also sufficiently modular and simple to allow PVP1 to be adapted to a given clinical context without altering function. Modular and well-documented code facilitates simple adaptation of the system to different hardware. Finally, we hope PVP1 can either directly or indirectly improve access to ventilators even beyond the COVID-19 pandemic, while also offering a reliable and open research platform for further ventilator development.

### Prior art and PVP1 Operation

The scientific community has mounted a powerful response to multiple facets of COVID-19, including numerous exciting ventilator projects, many of which leverage earlier designs developed to combat prior respiratory pandemics such as SARS and H1N1(1). In designing PVP1 we sought to learn from the challenges and limitations noted in prior studies while also providing a fully open source, transparent project. There are two key ventilator designs that received early FDA Emergency Use Authorizations–The University of Minnesota Ventilator (12), and the Mechanical Ventilator Milano (3). To handle production and FDA EUA approval, both projects eventually shifted manufacturer-of-record status to major companies–Boston Scientific, and Elemaster, respectively. Other academic projects such as the Vent4Us/PezGlobo ventilator (Stanford/University of Utah/University of Delaware) (13) have merged over time and also incorporated a variety of commercial backers. Still other projects such as the MIT E-Vent bag-valve ventilator design have remained open (www.emergency-vent.mit.edu) A more comprehensive discussion of numerous ventilator projects can be found in (1), where it is highlighted that the most successful projects have necessarily become less open due to constraints from industrial partners. Hence, a key goal with PVP1 was to describe and demonstrate a fully functional and modular ventilation platform that both highlights how effective a minimal design can be and provides a fully open platform for the broader community to leverage.

To accomplish this, PVP1 follows key FDA EUA design criteria by automating the classic Manley ventilator design (1, 10). The PVP1 schematic is shown in Figure 1A: the O2/air mixture is supplied to the system via a hospital gas blender, and the patient breath cycle is actively controlled via a proportional valve on the inspiratory limb and a solenoid valve with mechanical PEEP valve on the expiratory limb. Our approach integrates an embedded system (Raspberry Pi) supporting Pressure Controlled Ventilation (PCV), comprehensive monitoring of key alarm conditions, spontaneous breath detection (SIMV) and an intuitive touch-screen interface for clinician control. To use PVP1, the clinician programs a desired peak airway pressure (PIP), sets a manual PEEP-valve to establish expiratory pressure, and sets a target respiratory rate or I:E ratio. Convenient modifications to rise time and breath effort can be performed in real-time by the clinician. Core labeling specifications of PVP1 as required by the FDA EUA are presented in Table 1.

**Table 1.** PVP1 Specifications.

| Parameter | Range | Comments: tested range, theoretical performance, notes |
| --- | --- | --- |
| RR (BPM) | 10-40* | 12-20 tested based on ISO test tables; higher is feasible |
| TV (mL) | 200-500* | *Pressure controlled ventilation does not explicitly set TV; we validated resultant TV under PC as within the FDA EUA targets. |
| Flow Rate (L/min) | 0-100L/m | Valve specifications; maximum needed for testing was 85 L/m |
| PIP (cm H2O) | 15-60 | Tested up to 35 cm H2O during normal operation; safety hardware and alarms can support up to 60 cm H2O as per FDA EUA guidelines. |
| PEEP (cm H2O) | 5-25 | Validated PEEP range using approved commercial PEEP valves |
| I:E Ratios | 1:1-1:3 | Tested based on ISO 80601-2-80:2018 test tables |
| Available Ventilation Modes | PCV, SIMV | Modular system can be adapted for CPAP or non-invasive-modes at the software level. |
| Air Source | Hospital air | Rated for 50 psi, pre-blended oxygen/medical air mix. |
| Alarms | See Supplement | High/low airway pressure, hyper/hypoventilation, obstruction, low FiO2, PEEP not met, Disconnect/high leakage, Technical Alarms |
| Display variables | N/A | Airway pressure, expiratory flow, FiO2. Derived quantities: actual PIP, PEEP, VTE |
| Set variables | N/A | Target PIP, Target PEEP, Flow adjustment, Respiratory Rate, I:E ratio (or inspiratory time) |

## Results

Here, we present demonstrations and validations of how PVP1 performs key ventilatory processes. For clarity, we will first present results of using PVP1 (Core Performance), followed by a more detailed discussion of how PVP1 was designed at the hardware and software level. A complete description of the design process can be found at https://peoplesvent.org.

### Core Performance

#### Normal Operating Behavior

First, we evaluated the longterm stability and performance of PVP1 by performing standard pressure-controlled ventilation more than 70, 000 contiguous cycles over a period of 3 days (Fig. 2). All testing was performed using a high-grade test lung (Quicklung, Ingmar Medical) that offered the ability to tune compliance (C) and resistance (R) to meet FDA EUA test specifications (C=[5,20, 50] mL cm H2O; R=[5,20,50] cm H2O/L/s). Figure 2 shows pressure control performance for midpoint settings: C=20 mL/cm H2O, R=20 cm H2O/L/s, PIP=30 cm H2O, PEEP=5 cm H2O. PIP is reached within a 300 ms ramp period, then holds for the PIP plateau with minimal fluctuation of airway pressure for the remainder of the inspiratory cycle (blue). One the expiratory valve opens, exhalation begins and expiratory flow is measured (orange) as the airway pressure drops to PEEP and remains there for the rest of the PEEP period.

**Fig. 2.**
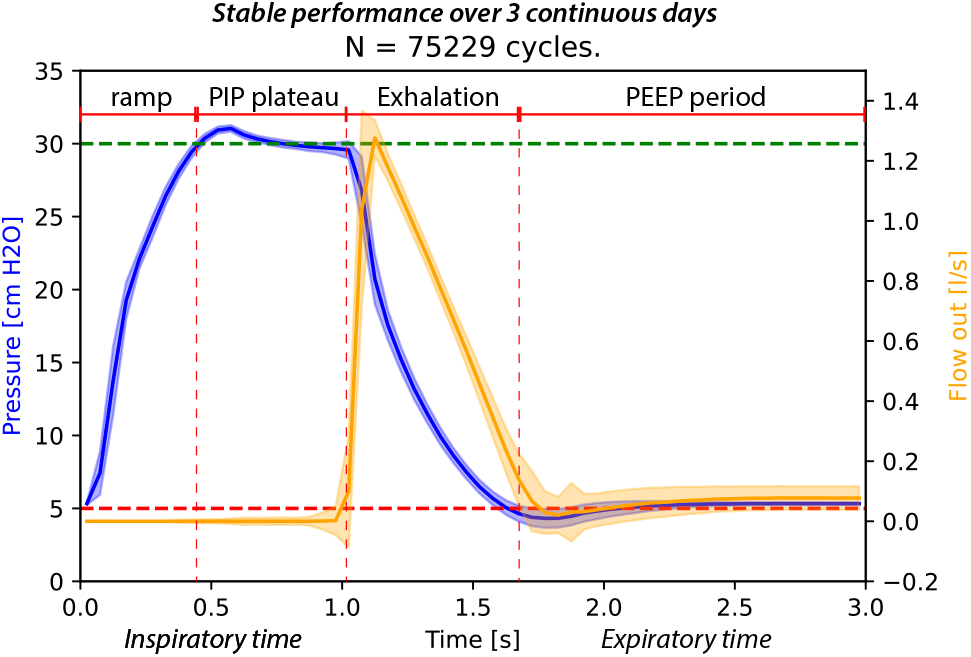
Overlaid pressure control breath cycle waveforms for airway pressure and flow out over 70,000+ cycles. Test settings: compliance C=20 mL/cm H2O, airway resistance R=20 cm H2O/L/s, PIP=30 cm H2O, PEEP=5 cm H2O.

Individual patient variation means that a one-size-fits all approach to pressure-controlled ventilation can have problems, and fine-tuning of key parameters such as the rise time (how quickly the ventilator reaches PIP) can allow more tailored ventilation. PVP1 supports such adjustment through a flow adjustment setting available to the clinician. This flow adjustment setting allows the user to increase the maximum flow rate during the ramp cycle to inflate lungs with higher compliance. The flow setting can be readily changed from the GUI and the control system immediately adapts to the user’s input. An example of this flow adjustment is shown in Figure 3A for four breath cycles. While all cycles reach PIP, the latter two have a higher mean airway pressure, which may be more desirable under certain conditions than the lower mean airway pressure of the former two.

**Fig. 3.**
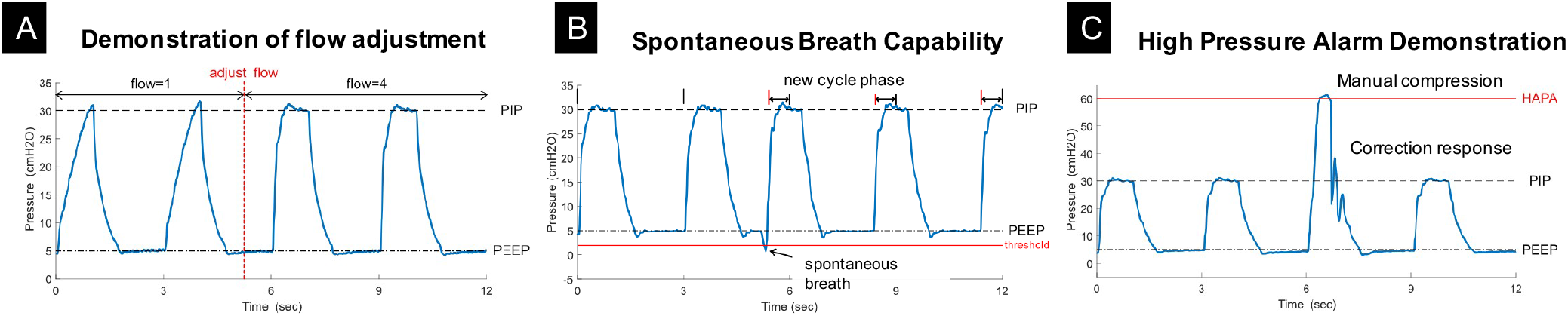
Demonstration of waveform tuning via flow adjustment. If necessary, the operator can increase the flow setting through the system GUI to decrease the pressure ramp time. Test settings: compliance C=20 mL/cm H2O, airway resistance R=20 cm H2O/L/s, PIP=30 cm H2O, PEEP=5 cm H2O.

#### Breath detection validation

A key feature of modern ventilators is to support spontaneous breath delivery should a non-anaesthetized patient try to breathe. Such patient-initiated breaths during the expiratory phase cause a sharp and transient drop in PEEP, and PVP1 can be set to detect these and trigger initiation of a new pressure-controlled breath cycle. We tested this functionality by triggering numerous breaths out of phase with the intended inspiratory cycle, using a device (QuickTrigger, IngMar Medical, Pittsburgh, PA) to momentarily open the test lung during PEEP and simulate this transient drop of pressure (Figure 3B).

#### Alarm response demonstration

Reliable and rapid alarm responses are a necessary feature of automated ventilators, and one of the most critical alarms (‘high priority’ in FDA EUA guidelines) for pressure control ventilation is the High-Airway-Pressure-Alarm (HAPA). According to peformance standards, the ventilator must detect abnormally high airway pressure must be detected and corrected within 2 breath cycles. In PVP1, the HAPA alarm can detect and respond to elevated airway pressure within 500 ms, while also throwing a high priority visual and audible alarm (Figure 3C).

#### EUA ISO Standard Tests

The FDA EUA guidelines lay out standardized tests required for all pressure controlled ventilation (see ISO 80601-20-80-2018). We performed this battery of tests (with the exception of those requiring controlled leak rates) and present the results in Figures 4 and 5. These tests cover an array of conditions, and more difficult test cases involve a high airway pressure coupled with a low lung compliance (case nos. 8 and 9 in Figure 5). Under these conditions, f the inspiratory flow rate during the ramp phase is too high, the high airway resistance will produce a transient spike in airway pressure which can greatly overshoot the PIP value. For this reason, the system uses a low initial flow setting and allows the clinican to increase the flow rate if necessary (as in Figure 3A).

**Fig. 4.**
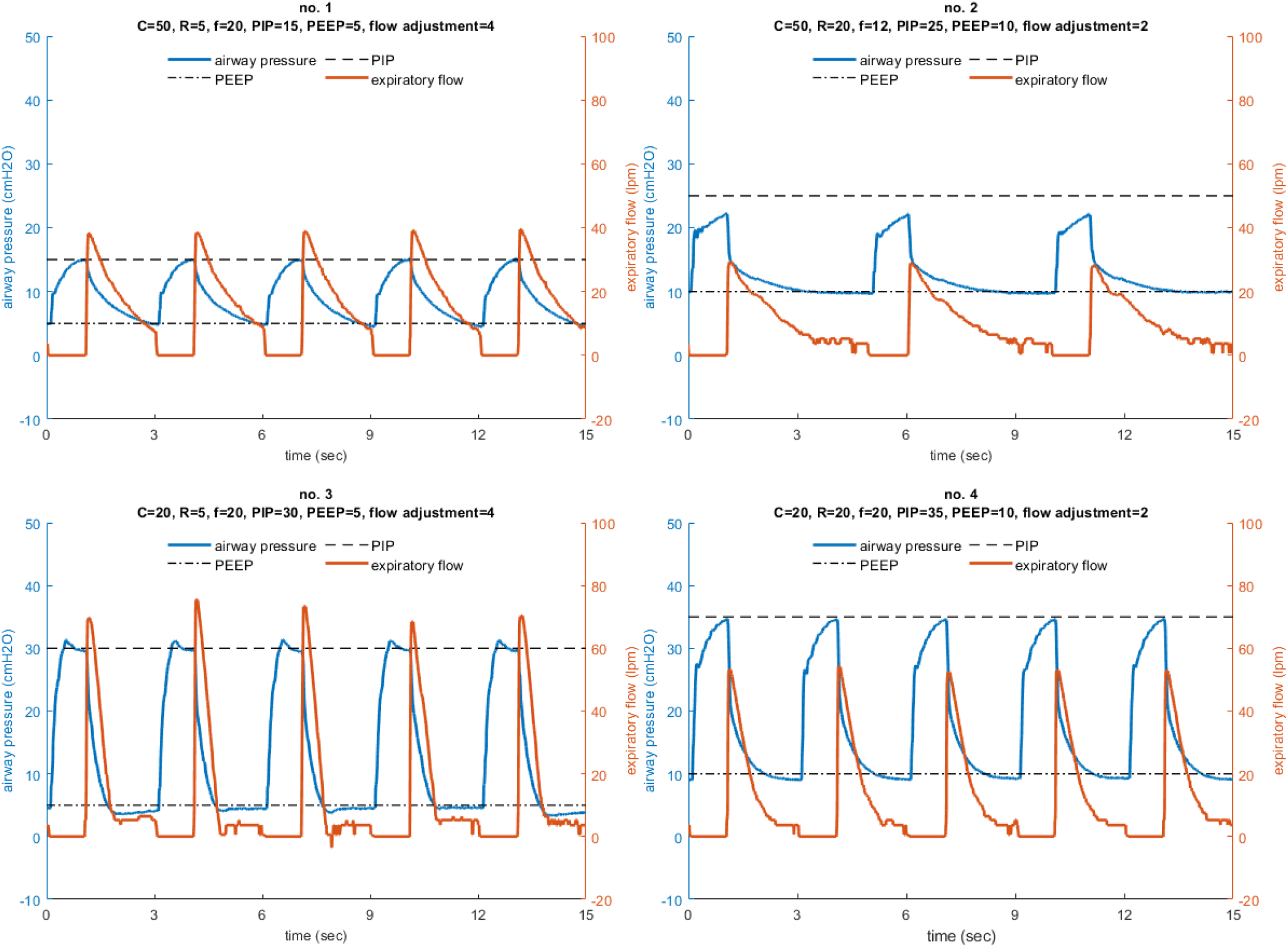
Performance results of the ISO 80601-2-80-2018 pressure controlled ventilator standard tests with an intended delivered tidal volume of 500 mL. For each configuration the following parameters are listed: the test number (from table 201.105 in the ISO standard), the compliance (C, mL/cm H2O), linear resistance (R, cm H2O/L/s), respiratory frequency (f, breaths/min), peak inspiratory pressure (PIP, cm H2O), positive end-expiratory pressure (PEEP, cm H2O), and flow adjustment setting. PIP is reached in every test condition except for case 2, which is approximately 2.4 cm H2O below the set point.

**Fig. 5.**
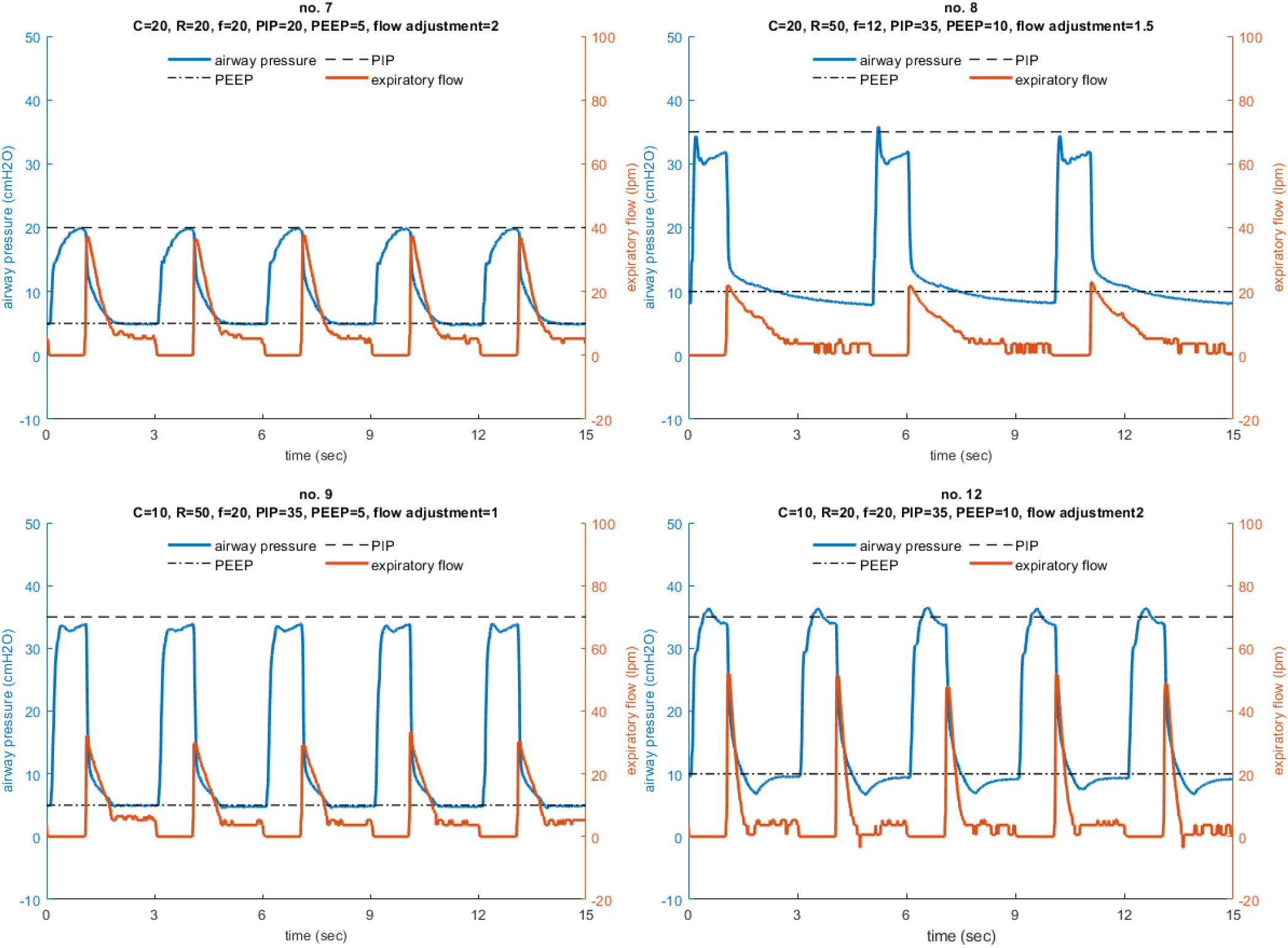
Performance results of the ISO 80601-2-80-2018 pressure controlled ventilator standard tests with an intended delivered tidal volume of 300 mL. For each configuration the following parameters are listed: the test number (from table 201.105 in the ISO standard), the compliance (C, mL/cm H2O), linear resistance (R, cm H2O/L/s), respiratory frequency (breaths/min), peak inspiratory pressure (PIP, cm H2O), positive end-expiratory pressure (PEEP, cm H2O), and flow adjustment setting. PIP is reached in every test condition.

The PVP1 integrates expiratory flow to monitor the tidal volume, which is not directly set in pressure controlled ventilation, but is an important parameter to ensure sufficient oxygen is delivered to the lungs. Of the test conditions in the ISO standard, four that we tested intended a nominal delivered tidal volume of 500 mL, three intended 300 mL, and one intended 200 mL. For most cases, the estimated tidal volume has a tight spread clustered within 20% of the intended value (see Figure 6).

**Fig. 6.**
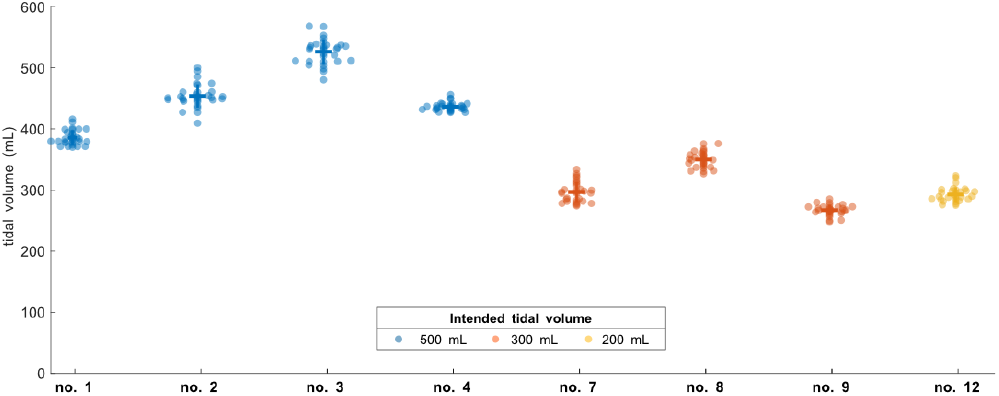
Tidal volume performance for the ISO 80601-2-80-2018 pressure controlled ventilator standard tests, averaged across 30 breath cycles for each condition.

The choice of items to display was done in close interaction with clinicians. Specifically, we chose to display VTE, Mean-airway-pressure (MAP), Peak-inspiratory-pressure (PIP), Positive end-expiratory pressure (PEEP), but the modular design of the GUI allows users to easily configure a different set of display and control values.

### Hardware design

The device components were selected to enable a minimalistic and relatively low-cost ventilator design, to avoid supply chain limitations, and to facilitate rapid and easy assembly. Most parts in our system are not medical-specific devices, and those that are specialized components are readily available and standardized across ventilator platforms, such as standard respiratory circuits and HEPA filters. We are providing a complete assembly of the device, including 3D-printable components, as well as justifications for selecting all actuators and sensors in the sections below, as guidance to those who cannot source an exact match to components used here. Readers may refer to the system schematic in Figure 7A.

**Fig. 7.**
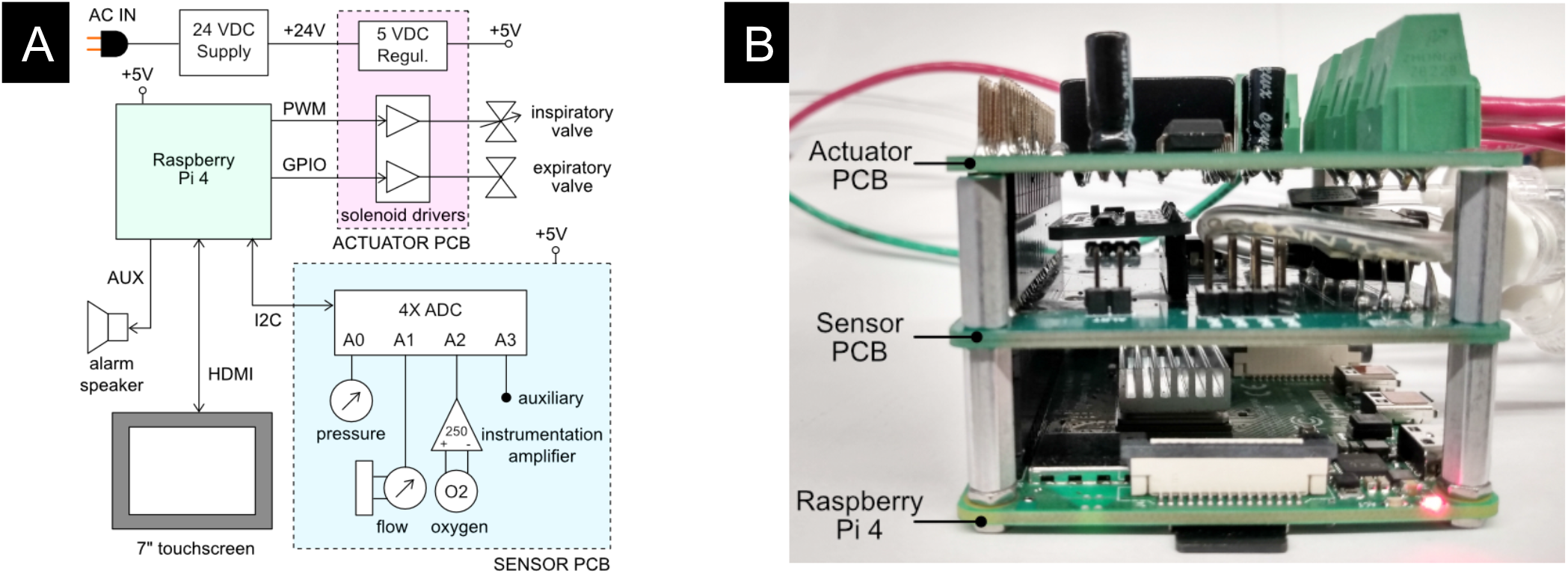
Overview of PVP1 electronics. A) Block diagram. System power is supplied by AC through an uninterruptible power supply (UPS) regulated to 24 VDC by a switched-mode power supply, to be used to drive the valves. This DC is further regulated to 5 VDC to supply power to the Raspberry Pi and associated sensors. The sensor PCB coordinates the inputs of the pressure and oxygen sensors. The actuator PCB amplifies control signals from the Pi to drive the valve solenoids. I/O is provided by a 7” touchscreen and an alarm speaker. B) Photograph of the modular system electronics. Two modular PCB ‘hats’ stack onto the Raspberry Pi via 40-pin stackable headers: the sensor PCB and the actuator PCB.

#### Hospital gas blender

At the inlet to the system, we assume the presence of a commercial-off-the-shelf (COTS) gas blender. These devices mix air from U.S. standard medical air and O2 as supplied at the hospital wall at a pressure of around 50 psig. The device outlet fitting may vary, but we assume a male O2 DISS fitting (NIST standard). In field hospitals, compressed air and O2 cylinders may be utilized in conjunction with a gas blender, or a low-cost Venturi-based gas blender. We additionally assume that the oxygen concentration of gas supplied by the blender can be manually adjusted. Users will be able to monitor the oxygen concentration level in real-time on the device GUI.

#### Fittings and 3D printed adapters

Standardized fittings were selected whenever possible to ease part sourcing in the event that engineers replicating the system need to swap out a component, possibly as the result of sourcing constraints within their local geographic area. Many fittings are American national pipe thread (NPT) standard, or conform to the respiratory circuit tubing standards (15mm I.D./22 mm O.D.). To reduce system complexity and sourcing requirements of specialized adapters, a number of connectors, brackets, and manifold are provided as 3D printable parts. All 3D printed components were print-tested on multiple 3D printers, including consumer-level devices produced by MakerBot, FlashForge, and Creality3D.

#### Pressure regulator

The fixed pressure regulator near the inlet of the system functions to step down the pressure supplied to the proportional valve to a safe and consistent set level of 50 psi. It is essential to preventing the over-pressurization of the system in the event of a pressure spike, eases the real-time control task, and ensures that downstream valves are operating within the acceptable range of flow conditions.

#### Proportional valve

The proportional valve is the first of two actuated components in the system. It enables regulation of the gas flow to the patient via the PID control framework, described in a following section. A proportional valve upstream of the respiratory circuit enables the controller to modify the inspiratory time, and does not present wear limitations like pinch-valves and other analogous flow-control devices. The normally closed configuration was selected to prevent over-pressurization of the lungs in the event of system failure.

#### Sensors

The system includes an oxygen sensor for monitoring oxygen concentration of the blended gas supplied to the patient, a pressure sensor located proximally to the patient mouth along the respiratory circuit, and a spirometer, consisting of a plastic housing (D-Lite, GE Healthcare) with an attached differential pressure sensor, to measure flow. Individual sensor selection will be described in more detail in a following section. The oxygen sensor read-out is used to adjust the manual gas blender and to trigger alarm states in the event of deviations from a setpoint. The proximal location of the primary pressure sensor was selected due to the choice of a pressure-based control strategy, specifically to ensure the most accurate pressure readings with respect to the patient’s lungs. Flow estimates from the single expiratory flow sensor are not directly used in the pressure-based control scheme, but enable the device to trigger appropriate alarm states in order to avoid deviations from the tidal volume of gas leaving the lungs during expiration. The device does not currently monitor gas temperature and humidity due to the use of an HME rather than a heated humidification system.

#### Pressure relief

A critical safety component is the pressure relief valve (alternatively called the “pressure release valve”, or “pressure safety valve”). The proportional valve is controlled to ensure that the pressure of the gas supplied to the patient never rises above a set maximum level. The relief valve acts as a backup safety mechanism and opens if the pressure exceeds a safe level, thereby dumping excess gas to atmosphere. Thus, the relief valve in this system is located between the proportional valve and the patient respiratory circuit. The pressure relief valve we source cracks at 1 psi (∼70 cm H2O).

#### Anti-suffocation check valve

A standard ventilator check valve (alternatively called a “one-way valve”) is used as a secondary safety component in-line between the proportional valve and the patient respiratory circuit. The check valve is oriented such that air can be pulled into the system in the event of system failure, but that air cannot flow outward through the valve. A standard respiratory circuit check valve is used because it is a low-cost, readily sourced device with low cracking pressure and sufficiently high valve flow coefficient (Cv).

#### Bacterial filters

A medical-grade electrostatic filter is placed on either end of the respiratory circuit. These function as protection against contamination of device internals and surroundings by pathogens and reduces the probability of the patient developing a hospital-acquired infection. The electrostatic filter presents low resistance to flow in the airway.

#### Standard respiratory circuit

The breathing circuit which connects the patient to the device is a standard respiratory circuit: the flexible, corrugated plastic tubing used in commercial ICU ventilators. Because this system assumes the use of an HME to maintain humidity levels of gas supplied to the patient, specialized heated tubing is not required.

#### HME

A Heat and Moisture Exchanger is placed proximal to the patient. This is used to passively humidify and warm air inspired by the patient. HMEs are the standard solution in the absence of a heated humidifier. While we evaluated the use of an HME/F which integrates a bacteriological/viral filter, use of an HME/F increased flow resistance and compromised pressure control.

#### Pressure sampling filter

Proximal airway pressure is sampled at a pressure port near the wye adapter, and measured by a pressure sensor on the sensor PCB. To protect the sensor and internals of the ventilator, an additional 0.2 micron bacterial/viral filter is placed in-line between the proximal airway sampling port and the pressure sensor. This is also a standard approach in many commercial ventilators.

#### Expiratory solenoid

The expiratory solenoid is the second of two actuated components in the system. When this valve is open, air bypasses the lungs, thereby enabling the lungs to de-pressurize upon expiration. When the valve is closed, the lungs may inflate or hold a fixed pressure, according to the control applied to the proportional valve. The expiratory flow control components must be selected to have a sufficiently high valve flow coefficient (Cv) to prevent obstruction upon expiration. This valve is also selected to be normally open, to enable the patient to expire in the event of system failure.

#### Manual PEEP valve

The PEEP valve is a component which maintains the positive end-expiratory pressure (PEEP) of the system above atmospheric pressure to promote gas exchange to the lungs. A typical COTS PEEP valve is a spring-based relief valve which exhausts when pressure within the airway exceeds a fixed limit. This limit is manually adjusted via compression of the spring. Various low-cost alternatives to a COTS mechanical PEEP valve exist, including the use of a simple water column, in the event that PEEP valves become challenging to source. We additionally provide a 3D printable PEEP valve alternative which utilizes a thin membrane, rather than a spring, to maintain PEEP.

### Actuator Selection

When planning actuator selection, it was necessary to consider the placement of the valves within the larger system. Initially, we anticipated sourcing a proportional valve to operate at very low pressures (0-50 cm H20) and sufficiently high flow (over 120 LPM) of gas within the airway. However, a low-pressure, high-flow regime proportional valve is far more expensive than a proportional valve which operates within high-pressure (∼50 psi), high-flow regimes. Thus, we designed the device such that the proportional valve would admit gas within the high-pressure regime and regulate air flow to the patient from the inspiratory airway limb.

Conceivably, it is possible to control the air flow to the patient with the proportional valve alone. However, we couple this actuator with a solenoid and PEEP valve to ensure robust control during PIP (peak inspiratory pressure) and PEEP hold, and to minimize the loss of O2-blended gas to the atmosphere, particularly during PIP hold.

#### Proportional valve sourcing

Despite designing the system such that the proportional valve could be sourced for operation within a normal inlet pressure regime (∼50 psi), it was necessary to search for a valve with a high enough valve flow coefficient (Cv) to admit sufficient gas to the patient. We sourced an SMC PVQ31-5G-23-01N valve with stainless steel body in the normally-closed configuration. This valve has a port size of 1/8” (Rc) and has previously been used in respiratory applications (reference?). Although the manufacturer does not supply Cv estimates, we empirically determined that this valve is able to flow sufficiently for the application (see Methods).

#### Expiratory solenoid sourcing

When sourcing the expiratory solenoid, it was necessary to choose a device with a sufficiently high valve flow coefficient (Cv) which could still actuate quickly enough to enable robust control of the gas flow. A reduced Cv in this portion of the circuit would restrict the ability of the patient to exhale. Initially, a number of control valves were sourced for their rapid switching speeds and empirically tested, as Cv estimates are often not provided by valve manufacturers. Ultimately, however, we selected a process valve in lieu of a control valve to ensure the device would flow sufficiently well, and the choice of valve did not present problems when implementing the control strategy. The SMC VXZ250HGB solenoid valve in the normally-open configuration was selected. The valve in particular was sourced partially due to its large port size (3/4” NPT). If an analogous solenoid with rapid switching speed and large Cv cannot be sourced, engineers replicating our device may consider the use of pneumatically actuated valves driven from air routed from a take-off downstream of the pressure regulator.

#### Manual PEEP valve sourcing

The PEEP valve is one of the few medical-specific COTS components in the device. The system configuration assumes the use of any ventilator-specific PEEP valve (Teleflex, CareFusion, etc.) coupled with an adapter to the standard 22 mm respiratory circuit tubing. In anticipation of potential supply chain limitations, as noted previously, we additionally provide the CAD models of a 3D printable PEEP valve.

### Sensor Selection

We selected a minimal set of sensors with analog outputs to keep the system design sufficiently adaptable. If there were a part shortage for a specific pressure sensor, for example, any readily available pressure sensor with an analog output could be substituted into the system following a simple adjustment in calibration in the controller. Our system uses three sensors: an oxygen sensor, an airway pressure sensor, and a flow sensor with availability for a fourth addition, all interfaced with the Raspberry Pi via a 4-channel ADC (Adafruit ADS1115) through an I2C connection.

#### Oxygen sensor

We selected an electrochemical oxygen sensor (Sensironics SS-12A) designed for the range of FiO2 used for standard ventilation and in other medical devices. The cell is self-powered, generating a small DC voltage (13-16 mV) that is linearly proportional to oxygen concentration. The output signal is amplified by an instrumentation amplifier interfacing the sensor with the Raspberry Pi controller (see Electronics Design section). This sensor is a wear part with a lifespan of ∼6 years under operation at ambient air; therefore under continuous ventilator operation with oxygen-enriched gas, it will need to be replaced more frequently. This part can be replaced with any other medical O2 sensor provided calibration is performed given that these parts are typically sold as raw sensors, with a 3-pin molex interface. Moreover, the sensor we specify is compatible with a range of medical O2 sensors, including the Analytical Industries PSR-11-917-M or the Puritan Bennett 4-072214-00, so we anticipate abundant sourcing options. The linearity of this sensor is ≤ 2% and the repeatability is ±1% volume O2 at 100% O2 applied for 5 minutes.

#### Pressure sensor (airway)

We selected a pressure sensor with a few key characteristics in mind: 1) the sensor had to be compatible with the 5V supply of the Raspberry Pi, 2) the sensor’s input pressure range had conform to the range of pressures possible in our device (up to 70 cm H2O, the pressure relief valve’s cutoff), and 3) the sensor’s response time had to be sufficiently fast. We selected the amplified middle pressure sensor from Amphenol (1 PSI-D-4V), which was readily available, with a measurement range up to 70 cm H2O and an analog output voltage span of 4 V. Moreover, the decision to utilize an analog sensor is convenient for engineers replicating the design, as new analog sensors can be swapped in without extensive code and electronics modifications, as in the case of I2C devices which require modifications to hardware addresses. Other pressure sensors from this Amphenol line can be used as replacements if necessary. The linearity of this sensor is ± 0.5% f.s., with 1% span shift across 5°C to 50°C.

#### Spirometer

Because flow measurement is essential for measuring tidal volume during pressure-controlled ventilation, medical flow sensor availability was extremely limited during the early stages of the 2020 COVID-19 pandemic, and supply is still an issue. For that reason, we looked for inexpensive, more easily sourced spirometers to use in our system. We used the GE D-Lite spirometer, which is a mass-produced part and has been used in hospitals for nearly 30 years. The D-Lite sensor is inserted in-line with the flow of gas on the expiratory limb, and two ports are used to measure the differential pressure drop resulting from flow through a narrow physical restriction. The third pressure-measurement port on the D-Lite is blocked by a male Luer cap, but this could be used as a backup pressure measurement port if desired. An Amphenol 5 INCH-D2-P4V-MINI was selected to measure the differential pressure across the two D-Lite take-offs. As with the primary (absolute) pressure sensor, this sensor was selected to conform to the voltage range of the Raspberry Pi, operate within a small pressure range, and have a sufficiently fast response time (partially as a function of the analog-to-digital converter). Also, this analog sensor can be readily replaced with a similar analog sensor without substantial code/electronics modifications. The linearity of this sensor is ±0.5% (maximum 0.25%) f.s., with 1% span shift across 5°C to 50°C.

### Electronics Design

The components of the PVP1 are coordinated by a Raspberry Pi 4 board, which runs the graphical user interface, administers the alarm system, monitors sensor values, and sends actuation commands to the valves (Figure 7). We elected to use a single Raspberry Pi rather than a computer connected to a dedicated microcontroller to minimize design complexity and to unify PVP1’s software in flexible, extensible, high-level Python modules. Dedicated microcontrollers are typically used because they run a single program without interruption, but they typically require ventilation control logic to be written in low-level C, and add an additional point of failure and inflexibility in the communication API between the computer and microcontroller. By taking advantage of the Raspberry Pi’s multiple processing cores and designing interfaces to a low-level hardware control daemon, PVP1 is capable of controlling ventilation at well below the latency of its hardware.

The main power to the systems is supplied by a DIN rail-mounted 150W 24V supply, which drives the inspiratory valve (4W) and expiratory valves (13W). This voltage is converted to 5V by a switched mode PCB-mounted regulated to power the Raspberry Pi and sensors. This power is transmitted across the PCBs through the stacked headers when required.

The core electrical system consists of two modular board ‘hats’, a sensor board and an actuator board, that stack onto the Raspberry Pi via 40-pin stackable headers (Figure 7B). The modularity of this system enables individual boards to be revised or modified to adapt to component substitutions if required.

#### Actuator Board

The purpose of the actuator board is twofold:

1. regulate the 24V power supply to 5V (CUI Inc PDQE15-Q24-S5-D DC-DC converter)
2. interface the Raspberry Pi with the inspiratory and expiratory valves through an array of solenoid drivers (ULN2003A Darlington transistor array)

#### Sensor Board

The sensor board interfaces four analog output sensors with the Raspberry Pi via I2C commands to a 12-bit 4-channel ADC (Adafruit ADS1015).

1. an airway pressure sensor (Amphenol 1 PSI-D-4V-MINI)
2. a differential pressure sensor (Amphenol 5 INCH-D2-P4V-MINI) to report the expiratory flow rate through a D-Lite spirometer
3. an oxygen sensor (Sensiron SS-12A) whose 13 mV differential output signal is amplified 250-fold by an instrumentation amplifier (Texas Instruments INA126)
4. a fourth auxiliary slot for an additional analog output sensor (unused)

A set of additional header pins allows for digital output sensors (such as the Sensiron SFM3300 flow sensor) to be interfaced with the Pi directly if desired.

Detailed schematics and board design files are included in the supplement.

### Software design

The software was modularly designed to facilitate future adaptation to new hardware configurations and ventilation modes. We carefully designed APIs for each of the modules to a) make them easily inspected and configured and b) make it clear to future developers how to adapt the system to their needs. The software has complete API-level documentation, making it so no part of the system is a black box.

All software development was done on GitHub, including a continuous-integration and automated testing suite with ≈ 97% code coverage and thorough code reviews of the core routines. All code was tested in parallel with the automated testing suite and on the physical device itself. The source code is publicly available on our GitHub repository, from which we also compile the documentation.

#### Software Architecture

The software is divided into two independent GUI and controller processes (Figure 8). The GUI process provides an interface to control and monitor ventilation, and the controller process handles the ventilation logic and interfaces with the hardware. Inter-process communication is mediated by a coordinator module via xml-rpc. Several ‘common’ modules facilitate system configuration and constitute the inter-process API. We designed the API around a unified, configurable `values` module that allow the GUI and controller to be reconfigured while also ensuring system robustness and simplicity.

**Fig. 8.**
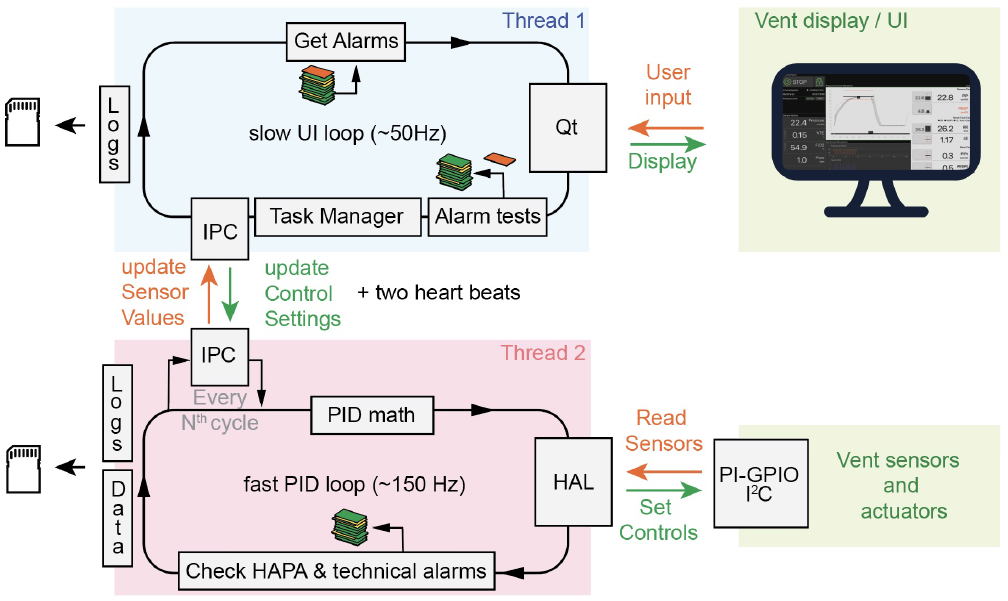
Overview of the software architecture. The user interface (UI) is controlled by a UI thread, that communicates with a second, faster, thread that implements the PID logic, by xml-rpc. The control loop is connected to hardware via the hardware abstraction layer (HAL). Both threads check for alarms; lower priority in the UI thread, and high priority in the controller thread, like HAPA (high airway pressure alert). The stack of cards illustrates a shared alarm-manager, which intelligently manages alarms, but keeping track of the various types, and sorting by importance. The individual processes add to this stack.

The multiprocess model has several key advantages over the single-process model:

1. **Robustness:** Since the GUI and controller are independent, a failing process can be restarted without interrupting the other.
2. **Efficiency:** Both processes can operate on their own processor core, allowing the controller to do rapid control operations without being interrupted by slower GUI operations.
3. **Flexibility:** Because the rapid controller process is separate from the slower GUI process, our system uses a single low-cost computer rather than a computer and an additional dedicated microprocessor. By using a single computer we were able to write the entire program in a single, high-level programming language and avoid the complexity induced by needing to design a separate controller firmware in low-level C. As such, it is easier to develop additional controller modules, GUI widgets, and hardware interfaces to implement different ventilation modes and hardware configurations in the future.

#### General software considerations

Across all software modules, we made sure to have a well-defined startup/stop sequence, to store all relevant raw data and measure the aging of components:

PVP1 should only be started, and stopped, using the provided user interface. Hard termination of the code, i.e. without allowing any more CPU time for executing commands, might freeze the valves in position, conceivably damaging the lung. It is therefore imperative to set valves into a safe positions, specifically, closing the inspiratory valve, to open the expiratory valve and thereby to relieve the system of any residual pressure and protect against future pressure-buildup. The same code has to be executed if any `Exception` occurs.

PVP1 continuously logs raw data into compressed hdf5 files. Pressure, the key variable of the controller, is sampled and stored at the speed of the controller main loop, while flow is only monitored during exhalation when the expiratory valve is open, and oxygen-concentration is measured every five seconds (see controller). In addition, we keep a log of events, as alarms and derived quantities and continuously produce human-readable logs as plain text that log overall program state and progress.

As hardware components age (e.g. the oxygen sensor and the valves), it is critical for the software to measure the time when it was first activated, and not just the number of breath cycles performed. To this end, the software stores a set of varbiables upon first activation, in the file `prefs.json` including the time of first start.

Core-load of the entire software package was ≈ 60%, and ≈ 40% for the two processes, and ≈ 15% for pigpiod, the demon that performs communication with the periphery. PVP1 thus puts the Pi’s ARM Cortex-A72 CPU with its four processor cores under only a mild load. Memory load was about 20% or 200 MB. The Raspberry Pi allows CPU frequency scaling, which enables the operating system to scale the CPU frequency up or down depending on demand. The governor, which regulates this scaling, was set to performance to deactivate this feature. In addition, stand-by and screensaver were deactivated.

#### Hardware I/O

The low-level firmware code is designed to be modular and makes extensive use of inheritance such that generic classes can be re-used for different applications, e.g. switching out a sensor, valve or ADC. In many cases, adapting the code to accommodate a new device requires re-writing a few lines of code.

We implement a hardware abstraction layer (HAL) to simplify the interaction between the controller and the firmware code so that all the complexity of the firmware code is hidden from the engineers who are developing the controls. Additionally, reconfiguration of the hardware is accomplished easily from the perspective of higher-level users due to the use of hardware config files.

For all communications over the GPIO pins of the Raspberry Pi, including sensor/ADC readings and valve control, we utilize the pigpio daemon (11). The pigpiod is a standalone interface which runs in its own process and is written in C. It is significantly faster than the default GPIO libraries and provides additional features like hardware-generated PWM signals. The pigpio Python library provides Python bindings for inter-process communication with the pigpio daemon.

Sensor reads require communication with pigpiod daemon which shares a single thread with the controller module. Future developers should strongly consider modifying the backend firmware in order to guarantee time-outs on communications. The sensors reading may occur in its own process such that the sensors are continuously sampled, with time-outs enabling the coordinator to re-start the daemon whenever it finds the daemon unresponsive.

#### Controller Module

Control into a breathing cycle was accomplished with a hybrid system of state and PID control. During inspiration, we actively control pressure using a PID cycle to set the inspiratory valve. Expiration was then instantiated by closing the inpiratory, and opening the expiratory valve to passively release PIP pressure as fast as possible. After reaching PEEP, we opened the inspiratory valve slightly to sustain PEEP using the aforementioned manually operated PEEP-valve and to sustain a gentle flow of air through the system.

Active control during inspiration was constructed around a single PID-style control loop. Pressure values were measured proximal to the patient, and communicated to the Raspberry PI using *I*^2^*C*. The Raspberry Pi compared this value against a user-provided target, and from that estimated proportional, derivative, and integral errors. Control coefficients were optimized manually on physical hardware by tuning against the set of EUA test conditions. The control-signal was then sent to the inspiratory and expiratory valves which caused a change in air-flow within ≈40 ms (related to the inductive load of the valve) (c.f. Figs. S3 and S4), while we ensure that the primary control loop ran at considerably higher speeds than this delay (see Methods).

We also allow the user to adjust flow through the system, by controlling `PIP_TIME` which is a multiplier for the PID coefficients. For lungs with higher compliance, more air is required to inflate to comparable pressures. As this assessment requires experience, we left the control of maximum flow rate to the clinician. During the primary control loop, pressure values have to be read as fast as possible. As the principal bottleneck is the communication with hardware, we chose to only read pressure values during inspiration. Flow out of the lung and oxygen concentrations are therefore only measured during expiration, where pressure and flow-readings alternate, and a single oxygen reading is obtained every five seconds.

In addition to pressure control, our software continuously monitors for autonomous breaths, high airway pressure, and general system status. Autonomous breathing was detected by transient pressure drops below PEEP. A detected breath triggered a new breath cycle. High airway pressure is defined as exceeding a threshold pressure for a minimum time (as to not be triggered by a cough). This triggers an alarm, and an immediate release of air to drop pressure to PEEP. The Controller also assesses whether numerical values are reasonable and updating over time (ie. the sensor hasn’t gotten stuck). If this is not the case, it raises an technical alarm. All alarms are collected and maintained by an intelligent alarm manager, that provides the UI with the alarms to display in order of their importance. Note that such smart designs, while not universally adopted, will be standard in the next generation of mechanical ventilators (6).

Measuring the expiratory flow, *F* (*t*) is sufficient to estimate VTE. To this end, we first estimated a baseline flow through the system, but not the lung (this is flow to sustain PEEP, see Control-section above). This was done by calculating a histogram of values in *F* (*t*) during expiration in a moving window, and applying a rank-filter to these numbers to define the baseline flow *F*_0_(*t*). We then integrated the difference over the expiration window *T* to obtain

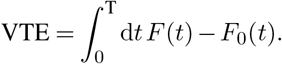

#### GUI Design

The GUI was designed to semantically segment system control, ventilation control, sensor monitoring, and alarm status while maintaining a unified syntax of interaction (Figure 9). We attempted to design the GUI so that it was intuitive enough to use without documentation by clearly separating and labeling the sections of the interface, providing few, clear points of control and interaction, and giving the user informative prompts to guide their use.

**Fig. 9.**
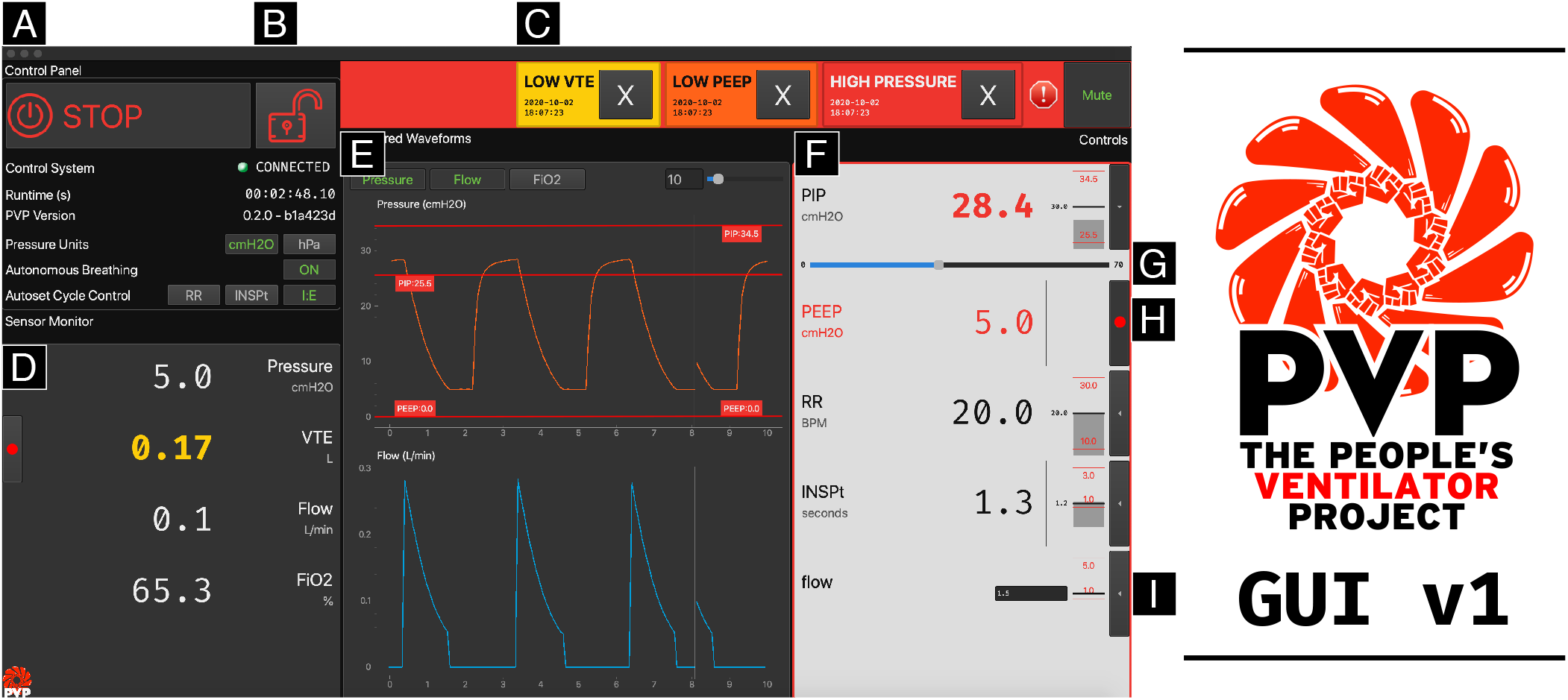
PVP1 GUI Design: The PVP1 GUI is composed of modular components to provide a uniform interaction syntax but also allow reconfiguration for different ventilation modes and hardware configurations. The GUI is broadly segmented into three columns: system monitoring components on the left (**A, D**), waveform plots of sensor values in the center (**E**), and ventilation controls on the right (**F**). A control panel (**A**) controls and displays basic system operating status. To prevent accidental changes, controls are locked (**B**) by default (but are unlocked in this figure). Beneath the control panel, a series of widgets in a “Monitor” column (**D**) display the values of sensors that are not used to control ventilation. The same widgets are to populate the rightmost “Control” column (**F**) to set the operating parameters of the ventilator. Ventilator control settings can be input with the touchscreen or mouse (with a slider, **G**), with the keyboard (in a text-entry box, **I**), or by recording recent sensor values (with a “record” button, **H**). By using a unified system of components across the whole interface, we can accomodate diverse hardware configurations (eg. rather than setting a computer-controlled `PEEP` in the interface, we use a mechanically controlled `PEEP` valve and set alarm thresholds by recording sensed values) and alarm conditions (`VTE` is a derived sensor value rather than a control value, but needs to have patient-calibrated alarm thresholds) while maintaining a consistent interaction syntax. Alarm thresholds are automatically calculated as some multiple of the set value (eg. a HAPA alarm is 115% of set `PIP`). Alarm thresholds are represented numerically and graphically in both the relevant control widget and waveform plot (red horizontal lines in **E** and **F**). Alarms are displayed as “Alarm Cards” (**C**) color coded and ordered by alarm severity. Color codes are also reflected in the relevant displayed parameter (eg. yellow displayed `VTE` value in **D** to match the low-severity `LOW VTE` alarm in **C**), allowing clinicians to quickly identify and attend to the cause of an alarm. Screenshot of actual GUI running PVP’s built in stimulation mode.

The GUI makes global system status clearly intelligible from across the room by using a uniform language of “alarm cards” (Fig. 9C) and color codes that indicate the severity and source of alarms to allow medical professionals to identify and address them as soon as possible. Alarm limits are displayed graphically next to the control widget and overlaid on relevant waveform plots (Fig. 9E, F) so values approaching a limit are clearly visible in either modality. The alarm system and sound design are described further in the supplemental materials (C).

Ventilation controls and sensor monitors (Fig. 9D, F) are displayed using a single widget class which makes them visually distinct but behave identically when setting alarm limits, displaying values, etc. These value display widgets support setting ventilation controls using the mouse/touchcreen (with a slider), keyboard, or by recording and averaging recent sensor values (Fig. 9G, H, I). All components of the GUI are modular and generated programmatically from the shared `values` module, which allows monitored and controlled values to be trivially reconfigured for different hardware configurations and ventilation modes while keeping a consistent API between the GUI and controller. The GUI also incorporates key considerations from the EUA, such as protection against accidental change and warnings against dangerous settings.

## Discussion

As described, PVP1 is a flexible, stable, and fully open platform for pressure controlled ventilation for a total cost of 1300 USD for low-volume production. With our combination of documentation, automated software tests, and modular design, this project is fully open source and offers anyone a state-of-the-art platform for exploring mechanical ventilation. Again, all necessary documentation and code are provided at the project website (peoplesvent.org). In the remainder of this section, we will discuss key areas for improvement and performance notes worth bearing in mind for those considering PVP1 for different use-cases.

### Overall Performance Assessment

PVP1 has demonstrated sustained operation over at least 70,000 continuous cycles without failure while maintaining stable ventilatory performance using a default test condition from the EUA test table.

PVP1 reaches PIP, and required VTE for key EUA tests, only deviating from the target PIP by 9% when ventilating with very high airway resistances (50 cmH2O/L/S; a challenging case for any ventilator). These are uncommon patient conditions and PVP1 performs even better in all other test cases. Future improvements to better compensate for challenging patient conditions could involve updates to the proportional valve to allow for finer control over flow, as well as more advanced control schemes to better modulate overshoot.

### Future Development Goals

PVP is intended to be a continually, communally developed project. PVP1 is released as a minimal implementation of a safe, invasive ventilator capable of Pressure Controlled Ventilation with spontaneous breath detection. There are, of course, many ways that the software and hardware design can be improved. Indeed its continual improvement is the point: we have developed and documented the system such that it is not a static design, but can be modified and improved as a general-purpose ventilation platform. We welcome programmers and users to submit issues to discuss bugs and needed developments, and submit their own improvements via pull requests.

#### Ventilation Modes

Modifications made purely at the software level would allow PVP1 to additionally support complete Pressure Supported Ventilation (PSV, for spontaneously breathing patients) as well as Non-Invasive Ventilation (such as Continuous Positive Airway Pressure, CPAP). At the hardware level, a valuable upgrade would be to incorporate an inspiratory flow sensor which would further open the possibility of Volume Controlled Ventilation and allow for the use of inspiratory flow for improved PSV. As PVP1 is inherently modular, these features can be added both to the open code base and to the assembly with little complication.

### I/O and Interfaces with other Systems

PVP1 is easy to integrate into existing software. Our data logger already supports export of all raw data into hdf5 and standard data formats (MatLab’s .mat and comma-separated-values .csv). A future version could automatically insert data into SQL-tables to facilitate integration into a patient’s file. Similarly, since our xml-rpc inter-process communication module operates over a network socket, it is straightforward to extend the software for centralized control or observation in hospital settings. We designed the software with a central loop time at least 10x faster than the valve-hardware delays. This can, in theory, allow the software to correct for hardware delays, and opens the door for future, improved control schemas.

## Disclaimer

PVP1 is not a regulated or clinically validated medical device. We have not yet performed testing for safety or efficacy on living organisms. All material described herein should be used at your own risk and does not represent a medical recommendation. PVP1 is currently recommended only for research purposes.

This document is not connected to, endorsed by, or representative of the view of Princeton University. Neither the authors nor Princeton University assume any liability or responsibility for any consequences, damages, or loss caused or alleged to be caused directly or indirectly for any action or inaction taken based on or made in reliance on the information or material discussed herein.

PVP1 is under continuous development and the information here may not be up to date, nor is any guarantee made as such. Neither the authors nor Princeton University are liable for any damage or loss related to the accuracy, completeness or timeliness of any information describe or linked to from this website.

## License Agreement

©2020: You may redistribute and modify this document, and related material, and make products using these under the terms of the CERN-OHL-S v2 (https:/cern.ch/cern-ohl). This source is distributed without any express or implied warranty, including of merchantability, satisfactory quality or fitness for a particular purpose. Please see the CERN-OHL-S v2 for applicable conditions.

## Data Availability

All necessary data are available at www.peoplesvent.org

## Acknowledgements

This work was supported by Princeton University which provided funding and facilities. JLS is supported by NSF Graduate Research Fellowship No. 1309047. We would also like to thank Grant Wallace, Zhenyu Song, Moritz Kütt and Philippe Bourrianne for very valuable discussions and technical support. In addition, we would like to thank Elad Hazan and Daniel Suo with the Google AI team at Princeton for testing the quality of our build instructions. We would also like to acknowledge the contribution of the open science community as a whole, by providing guidelines, standards and tools.

## Contributions

JL, CM, LS, DN, DJC conceptualized the project. DJC, JL, JLS, MS, TJZ wrote and edited the manuscript. **Hardware:** CM, JL, LS designed the hardware. TJZ designed the printed circuit boards. **Software:** MS developed the ventilation control system, the simulator and contributed to the core library, JLS developed the GUI, alarm system, and contributed to the core library, LS, JL developed the hardware interface and abstraction layer, MS, JLS developed the modular software architecture and automated tests. **Testing:** TJZ, MS, SD tested the deployed system and validated performance. JLS designed the alarm sounds, documentation website, and logo. DN provided medical advice. DJC managed the project and provided funding and resources.

## Supplemental Information

### A. Supplemental Materials and Methods

#### Calibration of the Flow Sensor

We calibrated the flow sensor using a mass flow controller (Alicat Scientific, MCR-100SLPM-D). The flow into the sensor was ramped up and then down in step of constant flow ranging from 10 L/min to 100 L/min (Figure S1 blue).

**Figure S1.**
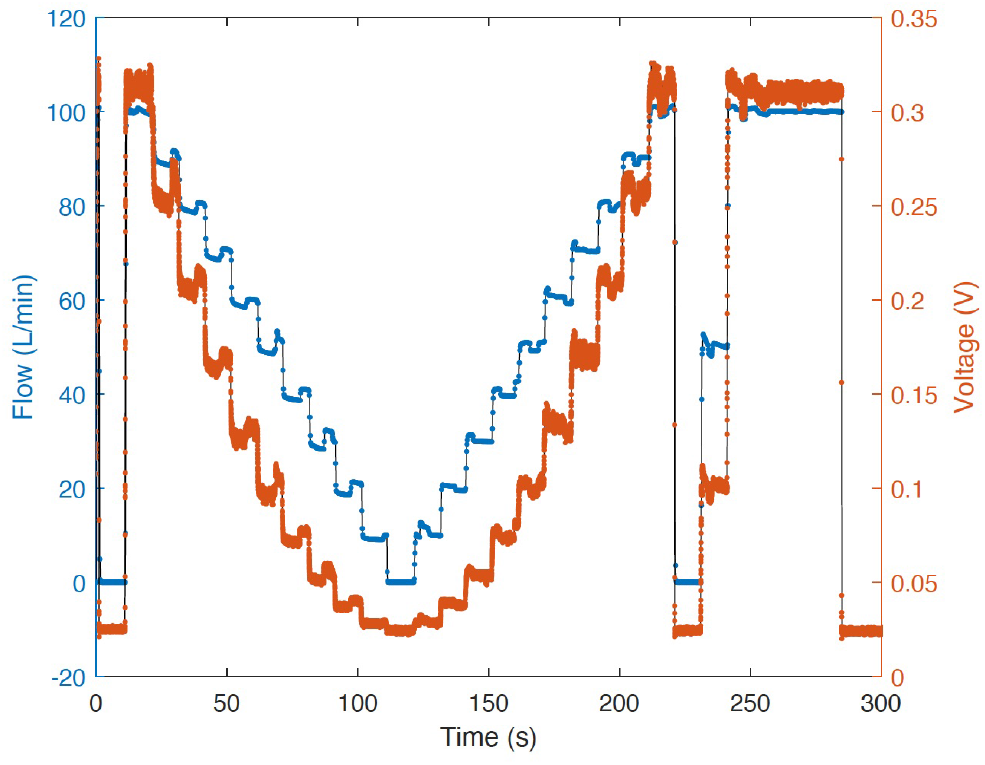
Calibration of the Flow Sensor. Flow introduced in steps of 10 L/min ranging from 10 L/min to 100 L/min. Blue points represent the flow measured by the mass flow controller and orange points represent the voltage recorded from the Dlite sensor.

At each step the flow was imposed for 10s and the voltage from the sensor was recorded (Figure S1 orange). Data points up to 5s after each jump and 1s before the next jump were discarded. For the remaining 4s the voltage data points were averaged to match the 10 Hz recording frequency of the mass flow controller. Subsequently both the flow and the voltage data was averaged at each step and used to calculate the calibration curve in Figure S2.

**Figure S2.**
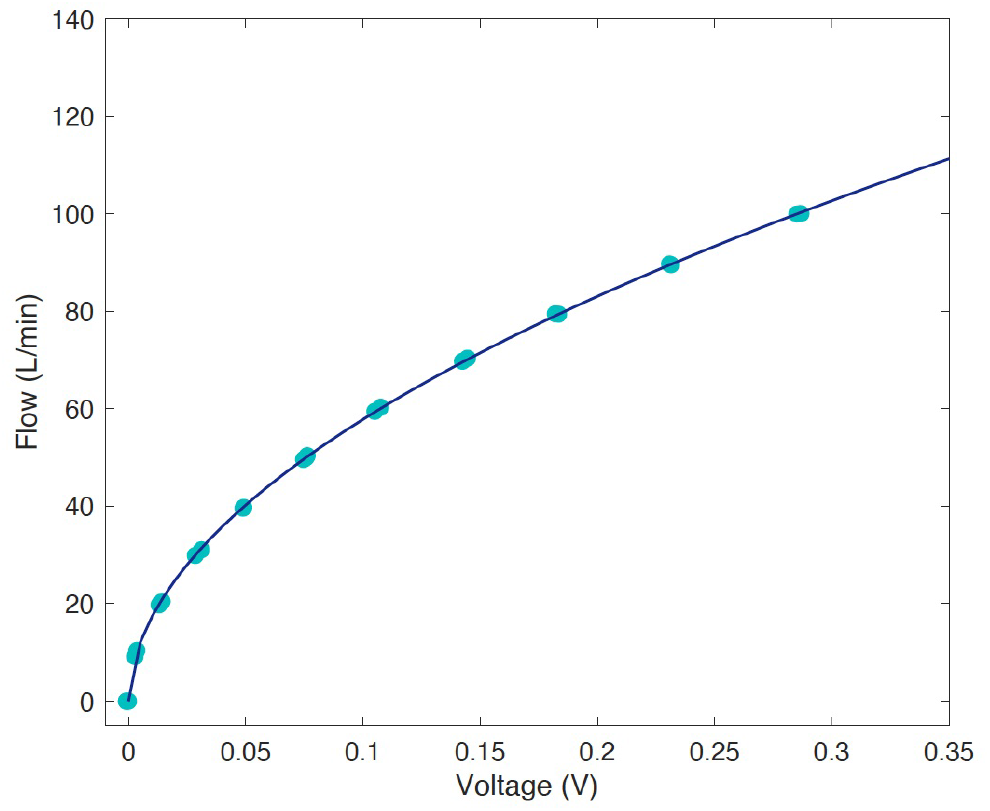
Calibration Curve for the Flow Sensor. Measurements of the flow as a function of the voltage recorded from the sensor. Turquoise points are the averaged data from the calibration and the blue line is the fit *V* = 192.6*F* ^1*/*1.91^.

To find the calibration curve the data was fit to a function of the form *V* = *a* · *F* ^1*/k*^ where the parameters were determined to be *a* = 192.6 and *k* = 1.91.

#### The Speed of the Control Loop

To measure the speed of the primary control loop, we saved the execution times of the control during while PVP1 was running with a lung simulator. The results are shown in Figure S3. Note the difference between inspiration and expiration; expiration is considerably slower, as additional sensor-reads for flow and oxygen sensing are required The Raspberry pi allowed for the primary control loop to run with a median loop-time of the entire software package of 6.6 ms during inspiration and 9.5 ms during expiration.

**Figure S3.**
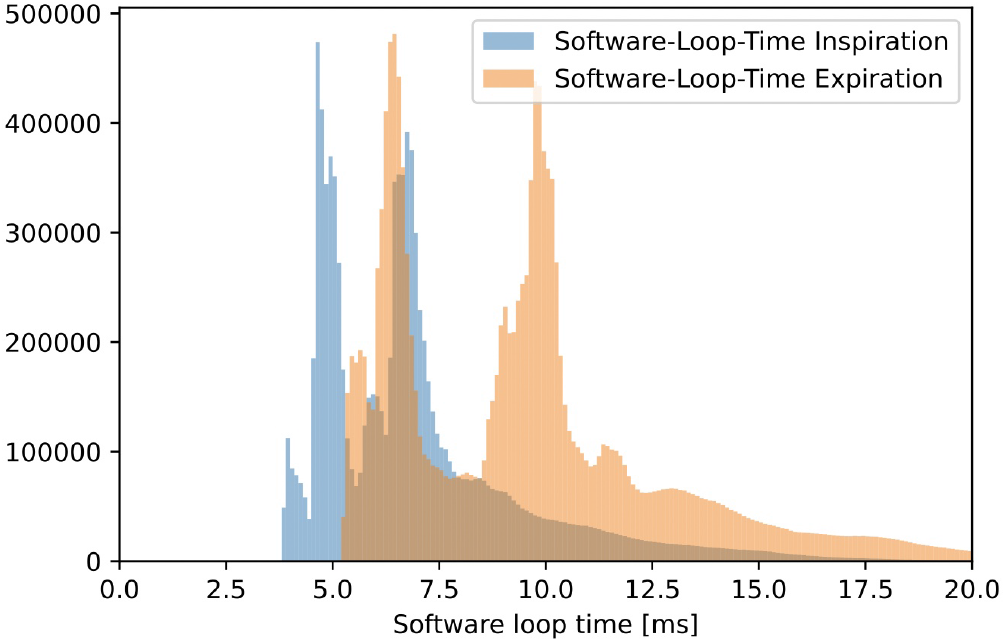
Histograms of the software-loop-time. Measurements of the software loop-time during inspiration (blue) and expiration (orange) over ca. 62h of breathing. Median inspiration loop time is 6.6 ms, median expiration loop time is 9.5 ms. Notice multiple peaks, depending on the addition of sensor readings, every one of which contributes ≈ 2.5 ms. The mechanical valves are a factor of ≈ 5 slower than the software.

#### Hardware delay characterization

To measure the delay of the inspiratory valve itself, we sent 100 consecutive ‘open’ and ‘close’ -commands to the inspiratory valve, while monitoring flow through the respiratory circuit.

Upon sending the control signal, flow increases with a delay of ≈ 50 ms. This is shown in Figure S4. We attribute this delay to the inductive load of the motor, and the finite time required for the mechanical opening of the valve. Note also, that this delay is considerably longer than the software-loop-time, and thus constitutes the principle bottleneck of the control system. This measurement was done with mild adjustments to the PVP1 software.

**Figure S4.**
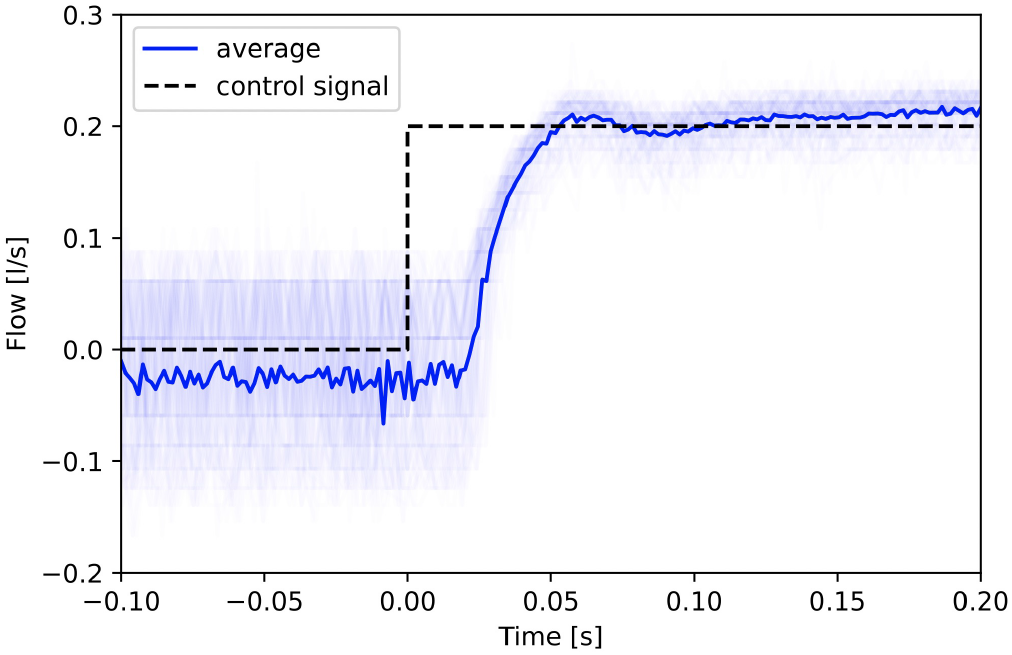
Measurements of the delay caused by the inspiratory valve. At time zero, the control signal to open the valve was sent and we monitored flow through the system. The thin blue lines illustrate 100 consecutive individual trials, the drawn blue line is the average, and the dashed line illustrates the control signal. 50% of the maximum flow was reached after 32 ms, 90% were reached after 53 ms.

### B. Supported Alarms

We support the following EUA alarms:

- Low Airway Pressure Alarm (LAPA) if target pressures (PIP or PEEP) is not reached, see Fig. 9.
- High Airway Pressure Alarm (HAPA) if pressure exceeds a critical limit, see Fig. 9.
- Hypoventilation Alarm if measured VTE is smaller than target, where target is defined in the first few breath cycles
- Tidal Volume not met Alarm if VTE is too small or too large.
- PEEP Alarm, if PEEP is not reached, see Fig. 9.
- Obstruction Alarm
- Disconnect / high leakage Alarm
- Oxygenation alarm, if the oxygen value deviates more than 5% from setpoint.
- Technical Alarm: A general class of alarms that are triggered whenever the software cannot work reliably.

### C. Alarm Design

Alarms are implemented as configurable `Alarm_Rule`s coordinated by a centralized `Alarm_Manger`. An alarm rule describes a) the `Condition`s for triggering an alarm, and b) the behavior and appearance of the alarm on the UI (Figure S5). Alarm conditions are implemented as composable classes that can accommodate complex triggering logic while remaining clear and inspectable (Figure S5, Lines 5-23).

#### Alarm Display

We try to balance salience of high severity alarms while minimizing unnecessary cognitive overhead by representing them as `Alarm_Card`s within an `Alarm_Bar` (Top of Figure 9). When no alarm is present, the bar is invisible, but takes the color of the highest-priority active alarm to give an unambiguous global status indicator. Alarm cards ensure each type of alarm is only represented once, allow individual control over dismissal/silencing of alarms, and visually triage lower-priority alarms by keeping them ordered by severity. The behavior of an alarm card is also determined by its alarm rule (See caption of Figure S5), so certain critical alarms aren’t missed but transient, lower-severity alarms don’t clutter the interface. The color-code of an alarm card is reflected in the widget that controls the relevant parameter (eg. the yellow `VTE` value in Figure 9), allowing attending physicians to quickly determine the source of alarms and how to correct them.

**Figure S5.**
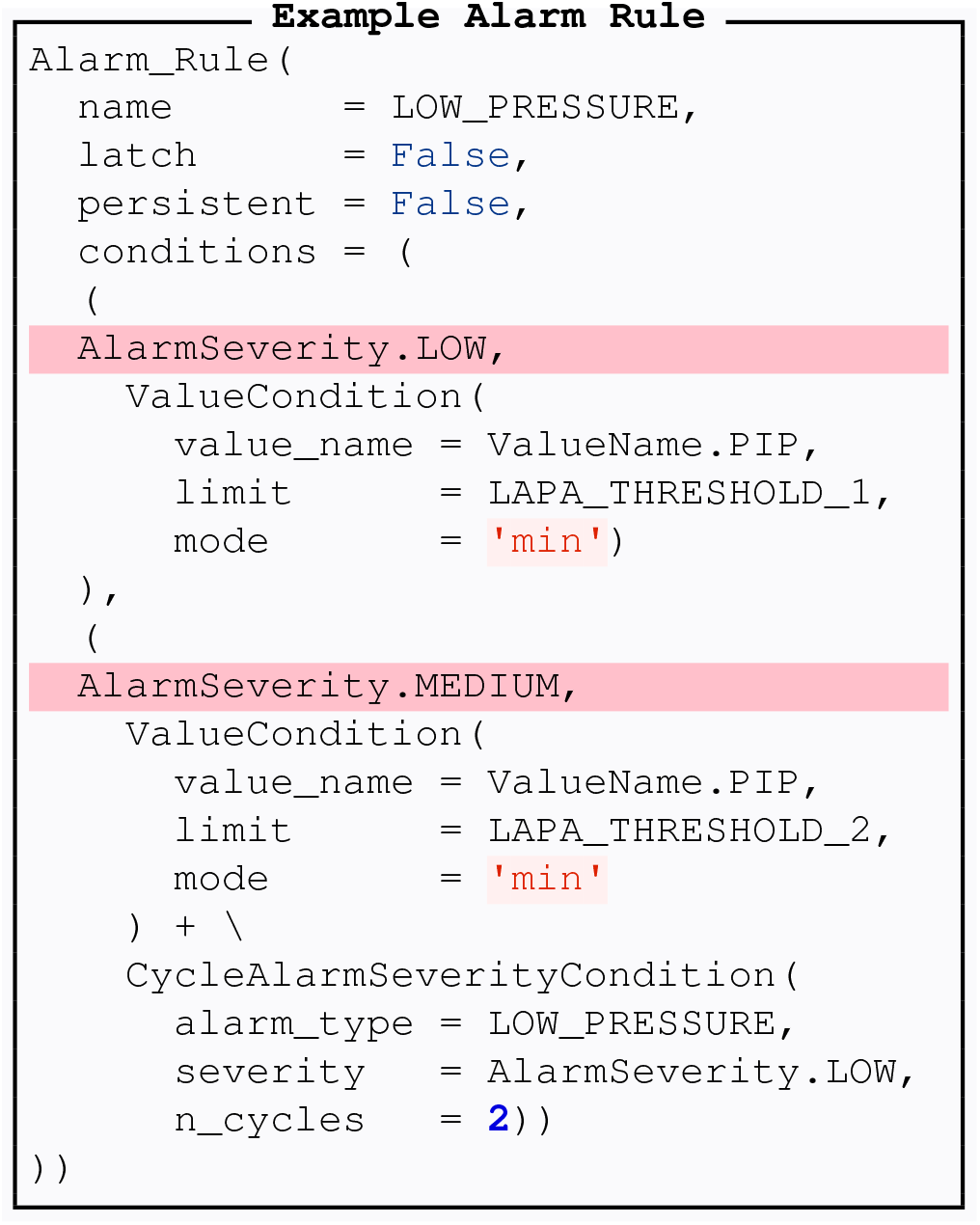
Example Low Pressure Alarm Rule. An Alarm Rule defines the behavior of an alarm in the GUI and the conditions for raising the alarm. Alarms can be `latch`ed (**L3**), where they cannot be visually dismissed until the alarm condition terminates or `persistent` (**L4**), where they will remain displayed until the user manually dismisses them. These settings ensure attending physicians never miss critical alarms, but are not overwhelmed with transient, low-severity alarms. Complex sets of `Condition`s for raising alarms can be described while remaining human-readable. This alarm has two severities (highlighted pink): a `LOW` severity alarm (**L7-11**) is triggered when `PIP` falls below some `LAPA_THRESHOLD`, which is escalated to a `MEDIUM` severity alarm (**L14-23**) if `PIP` falls below another threshold *and* the `LOW` severity alarm has been active for 2 breath cycles. Note how multiple conditions can be added (**L19**, literally with +) together, which allows triggering conditions to depend on multiple values, the states of other alarms, time, etc. In practice, rather than static alarm limits with a single value, all `Condition` values are updated from control values with some transformation (eg. this threshold could be kept at 15% below set `PIP`), but these dependencies have been omitted for brevity.

#### Alarm Sounds

We designed a set of alarm sounds (available in our repository) to be informative alert attending physicians while avoiding alarm fatigue. Only a single alarm sound is played at a time, and the alarm sound reflects the highest severity active alarm and the duration it has been active.

Alarm sounds are short (≈300ms) tone sequences, and severity of alarm is represented by pitch and the number of tones in each sequence - ie. a low-severity alarm is a repeating single low tone, and a high-severity alarm is a repeating sequence that adds two higher tones. At alarm onset, alarm sounds are low-pass filtered and have a lengthened attack and decay to soften their presentation while the physician first begins attending to the alarm condition. As alarms remain on, the filter, attack, and decay of the tone smoothly decrease to transition the sound to sharper, more urgent “clicks.”

All sounds use brief (≈40ms), noncontinuous tones that are silent for at least half of their duty cycle, leaving space for conversation and other sound. We attempt to disambiguate our alarms from other auditory alarms that could be present in the room by underlaying a soft pneumatic “sucking” sound synchronized to the tone sequences. Alarm sounds follow the same persistence rules as the relevant alarm rule (Figure S5, caption), reducing alarm fatigue by allowing transient, low-priority alarms to automatically silence themselves. Alarm sounds can also be muted entirely, or by dismissing specific alarms (“Mute” and “X” buttons in alarm bar in Figure 9, respectively) so they function to inform clinicians about patient state and then get out of the way.

### D. Supplemental Hardware Information

#### PCB Design

Printed circuit board schematics and bills of materials are presented here for the sensor PCB (Figure S7 and Table S1) and actuator PCB (Figure S6 and Table S2). PCB layout files are available on the project web page.

**Figure S6.**
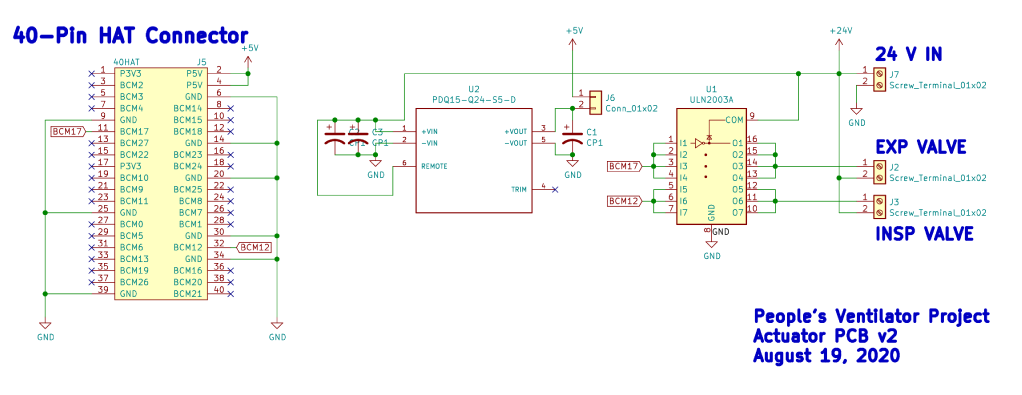
Actuator PCB schematic.

**Figure S7.**
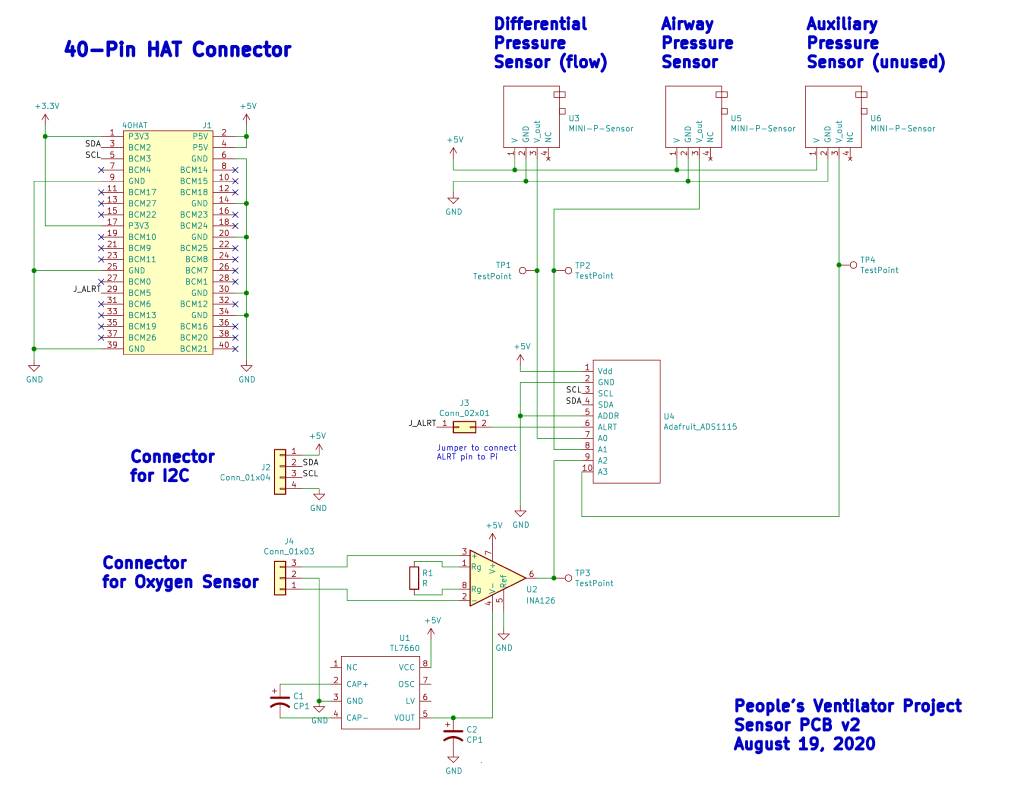
Sensor PCB schematic.

**Table S1.**
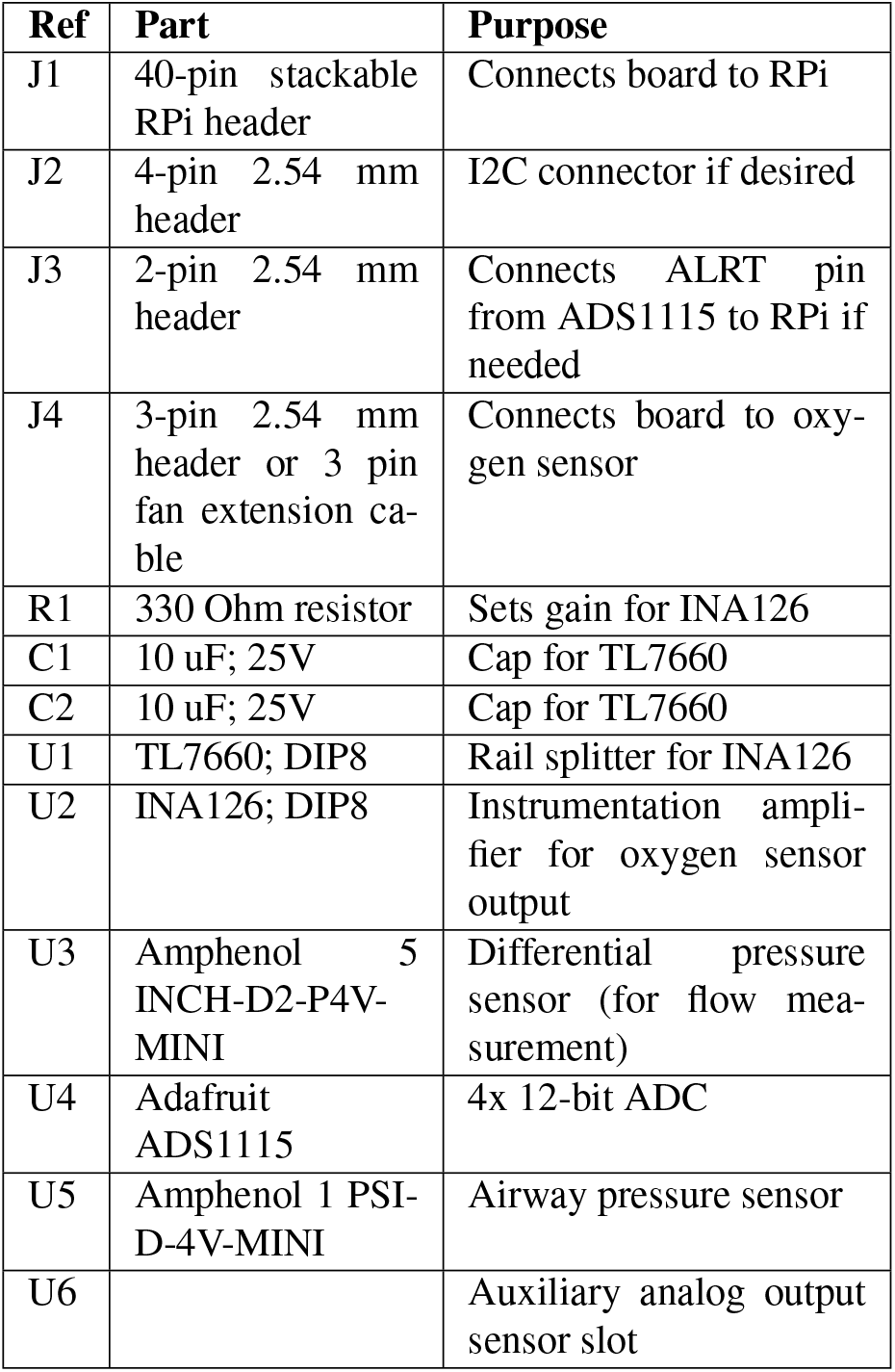
Sensor PCB bill of materials.

**Table S2.**
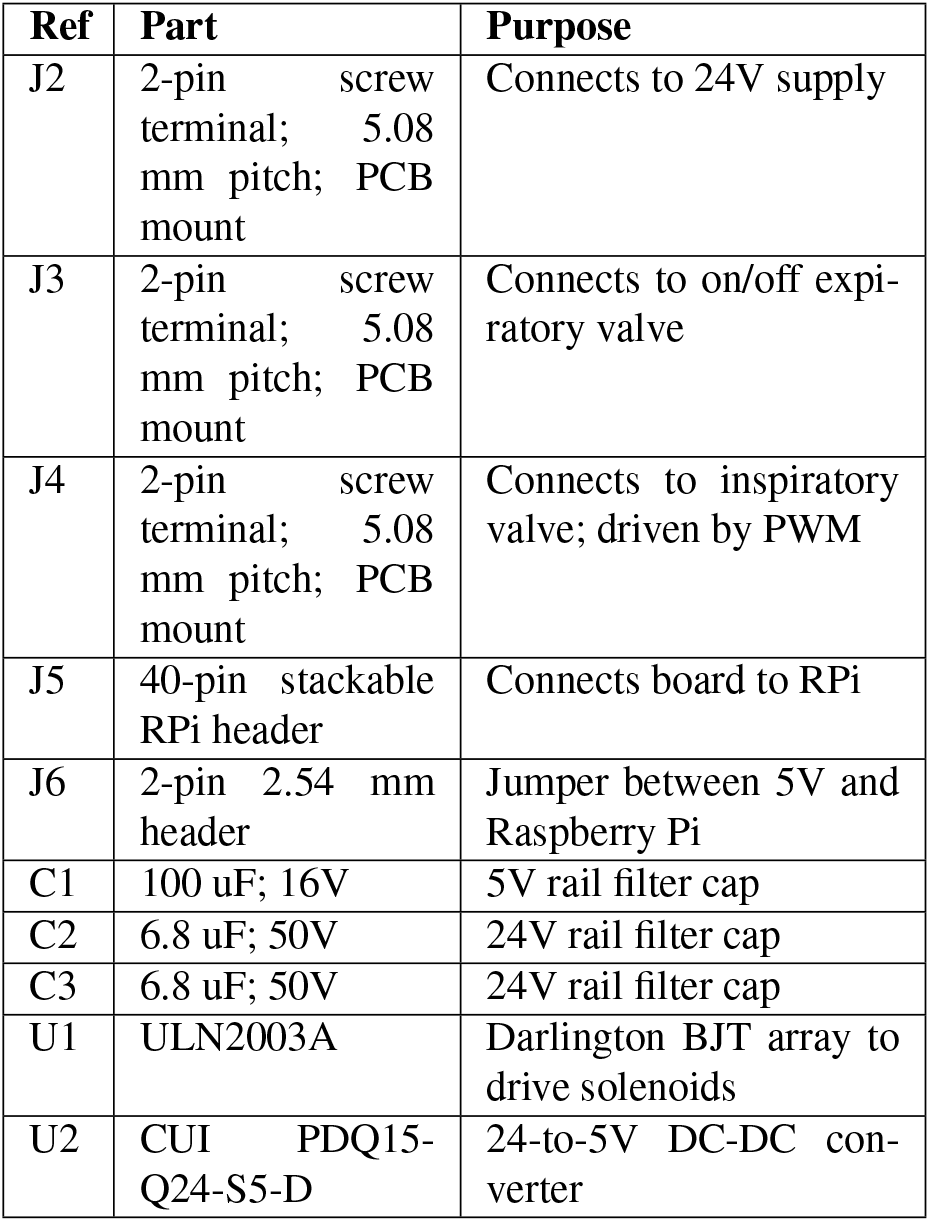
Actuator PCB bill of materials.

| Ref | Part | Purpose |
| --- | --- | --- |
| J2 | 2-pin screw terminal; 5.08 mm pitch; PCB mount | Connects to 24V supply |
| J3 | 2-pin screw terminal; 5.08 mm pitch; PCB mount | Connects to on/off expiratory valve |
| J4 | 2-pin screw terminal; 5.08 mm pitch; PCB mount | Connects to inspiratory valve; driven by PWM |
| J5 | 40-pin stackable RPi header | Connects board to RPi |
| J6 | 2-pin 2.54 mm header | Jumper between 5V and Raspberry Pi |
| C1 | 100 uF; 16V | 5V rail filter cap |
| C2 | 6.8 uF; 50V | 24V rail filter cap |
| C3 | 6.8 uF; 50V | 24V rail filter cap |
| U1 | ULN2003A | Darlington BJT array to drive solenoids |
| U2 | CUI PDQ15-Q24-S5-D | 24-to-5V DC-DC converter |

